# Cortical function and sensorimotor plasticity predict future low back pain after an acute episode: the UPWaRD prospective cohort study

**DOI:** 10.1101/2021.07.14.21260552

**Authors:** Luke C Jenkins, Wei-Ju Chang, Valentina Buscemi, Matthew Liston, Peter Humburg, Michael Nicholas, Thomas Graven-Nielsen, Paul W Hodges, James H McAuley, Siobhan M Schabrun

## Abstract

Predicting the development of chronic low back pain (LBP) at the time of an acute episode remains challenging. The Understanding persistent Pain Where it ResiDes (UPWaRD) study aimed to identify neurobiological and psychological risk factors for chronic LBP. Individuals with acute LBP (N=120) participated in a prospective cohort study with six-month follow-up. Candidate predictors were selected from the neurobiological (e.g. sensorimotor cortical excitability assessed by sensory and motor evoked potentials, Brain Derived Neurotrophic Factor genotype), psychological (e.g. depression and anxiety), symptom-related (e.g. LBP history) and demographic domains. Analyses involved multivariable linear regression models with pain intensity or disability degree as continuous variables. Secondary analyses involved a multivariable logistic model with presence of low back pain at six months (thresholding pain intensity and disability degree) as a dichotomous variable. Lower sensory cortex and corticomotor excitability, higher baseline pain intensity, higher depression, stress and pain catastrophizing were the strongest predictors (R^2^=0.47) of pain intensity at six months. Older age and higher pain catastrophizing were the strongest predictors (R^2^=0.30) of disability at six months. When LBP outcome was dichotomised, sensory cortex and corticomotor excitability, BDNF genotype, depression and anxiety, LBP history and baseline pain intensity, accurately discriminated those who did and did not report LBP at six months (c-statistic 0.91). This study identifies novel risk factors for future LBP after an acute episode that can predict an individual’s pain intensity and level of disability at six-month follow-up, and accurately discriminate between those who will and will not report LBP at six months.

## 1. INTRODUCTION

Low back pain (LBP) is the leading cause of years lived with disability worldwide [118] with approximately 40% of people continuing to experience pain for longer than 3 months (termed ‘chronic LBP’) [28]. Clinical strategies designed to ‘treat’ LBP once it has become chronic show at best, modest effect sizes regardless of intervention type [4;79;82;91]. An alternative strategy is secondary prevention, where the aim is to prevent the transition from acute to chronic pain. To achieve this goal, prognostic models that accurately discriminate between those who will, and will not, develop chronic LBP in the acute stage of pain are needed.

A limitation of current prognostic models is that biological, psychological and symptom-related risk factors are often studied in isolation, leaving predictive models that lack integration between psychological and symptom-related factors, and underlying biology [113]. Screening tools used in clinical practice to determine an individual’s risk of developing chronic LBP (e.g. STarT Back Screening Tool [44], and the short-form Orebro Musculoskeletal Pain Screening Questionnaire [69]) rely on self-report psychosocial and symptom-related factors. Although these tools allocate higher predicted risk scores to individuals who develop chronic pain, their ability to discriminate between those who will, and will not, develop chronic pain remains limited [54;55]. Prognostic models that integrate psychological (e.g. depression and coping strategies) and symptom-related factors (e.g. baseline pain intensity, history of prior LBP), explain up to 46% of the variance in LBP outcome [57]. Together these data suggest that although psychological and symptom-related factors predict the development of chronic LBP, a large proportion of variation in outcome is due to factors that are currently unmeasured or unknown [39;57]. The inclusion of neurobiological variables in prognostic models has the potential to predict a greater proportion of LBP outcome than currently possible [113].

Emerging evidence suggests several neurobiological risk factors with a putative link to LBP outcome that are yet to be evaluated in prognostic models. These include altered sensory and anterior cingulate cortex excitability [17;35], altered corticomotor excitability [19;94;109], Brain Derived Neurotrophic Factor (BDNF) genotype [10;22;59;98] and BDNF serum concentration [34;66]. These variables are relevant to LBP outcome as each has shown an association with the aberrant cortical plasticity thought to underpin chronic pain. Prior studies have shown altered excitability in the primary sensory (S1) and primary motor (M1) cortices that is associated with the development and maintenance of chronic pain [32;35;93]. A single nucleotide polymorphism in the human BDNF gene is associated with decreased behavioural driven changes in corticospinal output and cortical organization [9;20;34;37;59;83]. As serum BDNF concentration is associated with BDNF genotype [64] both measures are considered markers of neuroplastic potential [10;30].

The Understanding persistent Pain Where it ResiDes (UPWaRD) study aimed to determine whether neurobiological markers related to cortical function and plasticity (primary sensory cortex, anterior cingulate cortex and corticomotor excitability, BDNF genotype and serum concentration), psychological (anxiety and depression, pain catastrophizing, pain self-efficacy), symptom-related (baseline pain intensity, LBP history) and demographic (age, sex) risk factors assessed during an acute LBP episode could predict six-month LBP outcome.

## 2. METHODS

### 2.1. Study population

Details of the participants, recruitment and procedures for this cohort are described elsewhere [51]. In brief, 120 participants with an acute episode of LBP were recruited between 14 April 2015 and 23 January 2019 from local hospitals in South East Sydney and South Western Sydney Local Health Districts, New South Wales, Australia, primary care practitioners (e.g., general practitioners and physiotherapists), newspaper/online advertisements, flyers and social media sites. The date of the final participant’s six-month follow up was 25 July 2019.

Participants were eligible for inclusion if they had experienced acute LBP, reporting pain of at least 2/10 (Numerical rating scale [NRS], 0 = “no pain” and 10 = “worst pain imaginable”) at any time during the 7 days preceding initial screening [75]. Pain must have been present for more than 24 hours and less than six weeks duration following a period of at least one month without pain [29;75;100;120]. Acute LBP was defined as pain in the region of the lower back, superiorly bound by the thoracolumbar junction and inferiorly by the gluteal fold. Participants remained eligible if they had pain referred beyond this region that was not radicular pain from neural tissue involvement. At study inclusion, all participants with referred pain beyond the inferior gluteal fold underwent a physical examination by study staff to identify any sensory or motor deficit of the lower extremity. Participants with suspected lumbosacral radiculopathy characterised by the presence of weakness, loss of sensation, or loss of reflexes associated with a particular nerve root, or a combination of these, were excluded [13]. Any individual who presented with suspected serious spine pathology (e.g. fracture, tumour, cauda equina syndrome), other major diseases/disorders (e.g. schizophrenia, chronic renal disorder, multiple sclerosis), a history of spine surgery, any other chronic pain conditions or contraindications to the use of transcranial magnetic stimulation (TMS) were excluded [56]. Four assessors performed all study related procedures at laboratories located at Western Sydney University or Neuroscience Research Australia, Sydney, New South Wales, Australia. All procedures were approved by Western Sydney University (H10465) and Neuroscience Research Australia (SSA:16/002) Human Research Ethics Committees and conducted in accordance with the Declaration of the World Medical Association [5]. All participants gave written informed consent. Pre-planned methodology was published [51] and the trial registered (ACTRN12619000002189; Pre-results), adhering to recommendations of the PROGRESS initiative and TRIPOD statement [87;104].

### 2.2. Candidate predictors recorded at baseline

Fifteen candidate predictors were selected *‘a priori’* based on a theoretical association with the development of chronic pain and supporting evidence from cross-sectional studies [10;35;40;68;92;109]. Justification for each variable and specific methodology is provided in the study protocol [51]. In brief:

#### 2.2.1. Sensory and anterior cingulate cortex excitability

Participants were seated comfortably in a chair with feet on the floor and arms relaxed. Participants were asked to sit still, keep their eyes closed and not to fall asleep for the duration of the test procedure. A bipolar electrode (silver-silver chloride disposable electrodes, inter-electrode distance 2.0 cm; Noraxon USA, Arizona, USA) was positioned 3 cm lateral to the L3 spinous process, ipsilateral to the side of the worst LBP and a constant current stimulator (Digitimer Ltd, Hertfordshire, UK DS7AH) delivered non-noxious electrical stimuli (single stimulus; pulse width 1 ms). The anode was the inferior attachment of the bipolar electrode. Electrical stimulation was increased in 1 mA increments until the perceptual threshold was reached. The testing intensity was set at three times the perceptual threshold. If this testing intensity evoked pain, it was decreased in 1 mA increments until the stimulus was reported as non-noxious.

Sensory evoked potentials (SEPs) were recorded in response to two blocks of 500 non-noxious electrical stimuli (∼2 Hz, with random interval schedule of 20% to decrease accommodation), using gold plated cup electrodes positioned over S1 (3 cm lateral and 2 cm posterior of Cz) on the hemisphere contralateral to the side of worst LBP and referenced to Fz using the International 10/20 System [46]. The side of worst LBP was determined on the day of baseline testing by asking the participant “on average over the past 24 hours which side of your back is most painful?” If the participant was unable to determine the most painful side and reported central LBP over the previous 24 hours, SEPs were recorded on the hemisphere contralateral to the dominant hand. Electrode impedance was maintained at <5 k*Ω*. EEG signals were amplified 50,000 times, band pass filtered between 5-500 Hz, and sampled at 1,000 Hz using a Micro1401 data acquisition system and Signal software (CED Limited, Cambridge, UK).

Individual SEP traces were inspected and those with eye movements, muscle artefacts or electrical interference were rejected. Less than 15% of all SEP traces were excluded. Remaining traces from the two SEP blocks were averaged for each participant and used for analysis [18]. The averaged wave form was full-wave rectified and the area under the rectified curve (µV) determined for the N_80_ (between the first major downward deflection of the curve after stimulus onset and the first peak, N_80_), N_150_ (between the first and second peak, N_80_ and N_150_, respectively) and P_260_ (between the second negative peak, N_150_ and the positive deflection of the curve starting around 150 ms after stimulus onset, P_260_) time epochs [32]. **Figure 1** displays a rectified trace with time epochs used for analysis. The SEP area measurement was chosen for analysis as it is less susceptible to signal-to-noise ratio concerns [24], and considered superior to peak-based measures for assessing event-related potentials [32;53;84;121]. Previous electrophysiological research suggests distinct SEP components reflect sensory afferent processing within discrete brain regions [7] - the N_80_ SEP time epoch is thought to reflect processing in S1, the N_150_ SEP time epoch is thought to reflect processing in the secondary sensory cortex (S2) and the P_260_ time epoch thought to reflect processing within the anterior cingulate cortex (ACC)[32;35].

**Figure 1.**
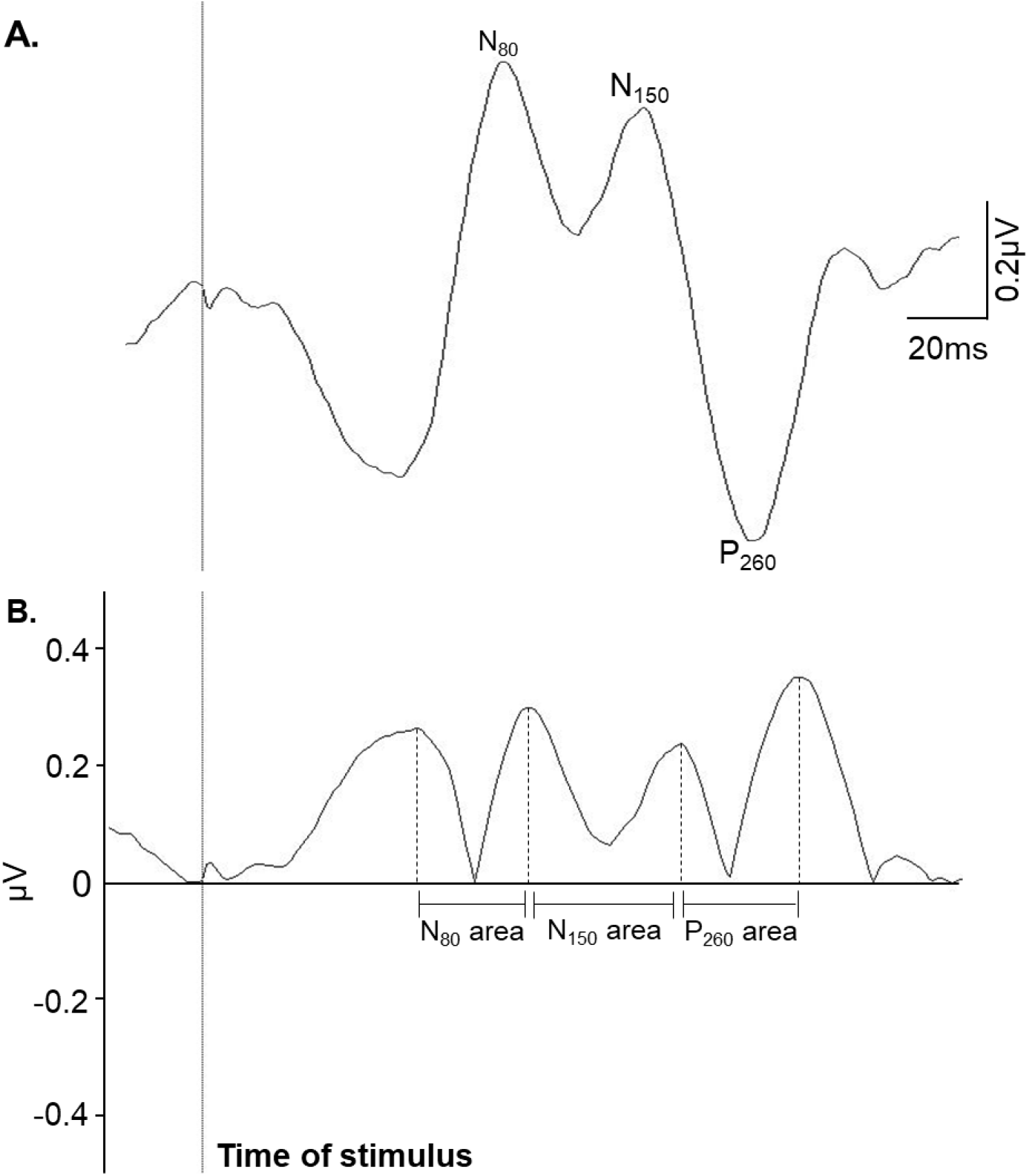
Example of a sensory evoked potential (SEP) recorded from the paraspinal muscles (average of 500 traces) from a single participant. **Figure 1A** shows the N_80_, N_150_ and P_260_ SEP peaks. **Figure 1B** shows the area under the rectified curve for each time epoch (N_80_, N_150_ and P_260_ area) that was used for analysis.

#### 2.2.2. Corticomotor excitability

Participants underwent a standardised TMS mapping procedure [78;95;109]. Single-pulse, monophasic stimuli (Magstim 200 stimulator, 7 cm figure-of-eight coil; Magstim Co. Ltd. Dyfed, UK) were delivered over the primary motor cortex (M1) contralateral to the side of the worst LBP. The coil was positioned tangential to the skull with the handle pointing posterior-laterally at 45 degrees from midline [11;52;74]. Participants wore a cap marked with a 6 x 7 cm grid oriented to the vertex (point 0, 0). The vertex was determined using the International 10/20 System, and aligned with the centre of the cap [43]. The cap was tightly fitted and the position regularly checked to ensure placement consistency. Starting at the vertex, five stimuli were delivered over each site on the grid with an inter-stimulus interval of 6 s. Electromyography (EMG) was recorded from the paraspinal muscles 3 cm lateral to the spinous process of L3 and 1 cm lateral to the spinous process of L5 on the side of the worst LBP using disposable Ag/AgCl surface electrodes (Noraxon USA Inc, Arizona, USA) [65;77]. Ground electrodes were placed over the anterior superior iliac spine bilaterally. EMG data were amplified 1000x, filtered between 20-1000 Hz and sampled at 2000 Hz using a Micro1401 data acquisition system and Spike2 software (CED Limited, Cambridge, UK). As 120% of active motor threshold often exceeds the maximum stimulator output [109], all stimuli were delivered at 100% of stimulator output while participants activated the paraspinal muscles to 20 ± 5% of their EMG recorded during a maximum voluntary contraction (MVC; determined as 20% of the highest root mean square [RMS] EMG averaged over 1 s during three, 3-s maximal muscle contractions performed against manual resistance in sitting) [94;96;109]. Real-time feedback of time paraspinal muscle RMS EMG and the target level was displayed on a monitor for the duration of the test procedure [111]. All TMS procedures adhered to the TMS checklist for methodological quality [21].

TMS motor evoked potentials (MEP) were analysed using MATLAB 2019a (The MathWorks, USA). Onset and offset of the MEPs in each individual trace were visually identified then averaged at each scalp site. The amplitude of paraspinal MEPs was measured as the RMS EMG amplitude, between the onset and offset of the MEP from which the background RMS EMG was subtracted (RMS amplitude recorded between 55–5 ms prior to stimulation) [109]. For frames with no visually identifiable MEP onset and offset, RMS EMG amplitude between a 20-to 50-ms time window after stimulation was extracted [16]. Paraspinal MEP amplitudes were normalized to the largest MEP amplitude across sites and superimposed over the respective scalp sites to generate a topographical map. A scalp site was considered active if the normalised MEP amplitude was equal to or greater than 25% of the peak response [18]. Normalised values below 25% of the peak response were removed and the remaining values rescaled from 0 to 100% [95;110].

Two parameters were calculated from the normalised motor cortical maps. First, L3 and L5 map volumes were calculated as the sum of normalized MEP amplitudes recorded at all active scalp sites [119]. Second, the centre of gravity (CoG), defined as the amplitude weighted centre of the map, was calculated for the M1 cortical representation of L3 and L5 paraspinal muscles using the formula: CoG = ΣVi · Xi/ΣVi, ΣVi · Yi/ΣVi where: Vi=mean MEP response at each site with the coordinates Xi, Yi [112;119]. Distance between the L3 and L5 CoG (L3/L5 CoG overlap) was calculated as the Euclidean distance (ED) using the formula: ED = √((Y_L3_ – Y_L5_)^2^ + (X_L3_ – X_L5_)^2^), where Y = anterior-posterior coordinates; X = medial-lateral coordinates of L3 and L5 [116;117].

#### 2.2.3. BDNF genotyping

Buccal swabs were taken on the day of baseline testing (Isohelix DNA Isolation Kit) [22]. Samples were immediately frozen and stored at -80°C. Genomic DNA samples were polymerase chain reaction amplified and sequenced by the Australian Genome Research Facility (AGRF). Genotyping was performed as recommended by the manufacturer with reagents included in the iPLEX Gold SNP genotyping kit (Agena) and the software and equipment provided with the MassARRAY platform (Agena) [23]. Consistent with prior investigations [10;63;73], the BDNF gene was coded as a dichotomous variable (AA/AG or GG). The more common G allele encodes Valine (Val), while the A allele encodes Methionine (Met).

#### 2.2.4. BDNF serum concentration

Peripheral venous blood was drawn into serum tubes (BD, SST II Advance) through venepuncture of the median cubital vein at baseline. The sample was clotted (30 min, room temperature) then separated by centrifugation (2500 rpm, 15 min). The samples were pipetted into 50 μL aliquots and stored at -80°C. After thawing, the Simple plex Ella™ platform was used to analyse the specific expression of BDNF. Briefly, 10 μL samples were diluted with 90 μL of sample diluent then added to the cartridge, according to the standard procedure provided by the manufacturers (Protein Simple, CA, USA). All steps in the immunoassay procedure were carried out automatically and scans were processed with no user activity. Cartridges included built-in lot–specific standard curves. Single data (pg/mL) for each sample were automatically calculated and converted to ng/mL for statistical analysis. The limit of detection was 5.25 pg/mL. Fifteen randomly selected samples were analysed in duplicate and demonstrated near perfect correlation (r = 0.98, *P* = < 0.001).

#### 2.2.5. Psychological status

Three questionnaires were used to assess specific aspects of psychological status known to be of relevance to the development of chronic LBP: depression and anxiety [15], pain catastrophising [14] and pain beliefs [67]. The 21-Item Depression, Anxiety and Stress Scales Questionnaire (DASS-21) was used to assess depression, anxiety and stress. A total score between 0 and 63 was calculated, where higher scores indicate higher levels of depression, anxiety and/or stress [3;80]. The 13-item Pain Catastrophizing Scale (PCS) assesses distressing thoughts related to painful experiences [105]. A total score between 0 and 52 was calculated, where higher scores represent more severe catastrophic thoughts about pain [105]. The 10-item Pain Self-Efficacy Questionnaire (PSEQ) was used to assess an individual’s beliefs in their ability to perform a range of functional activities while in pain. A total score between 0 and 60 was calculated, where higher scores represent greater self-efficacy beliefs [76]. The DASS-21 (Cronbach’s α = 0.88), PCS (Cronbach’s α = 0.87) and PSEQ (Cronbach’s α = 0.92) have all demonstrated high degrees of reliability (internal consistency) [41;76;105].

#### 2.2.6. Symptom-related factors

Baseline pain intensity was drawn from the Brief Pain Inventory (BPI) administered on the day of baseline testing. Participants rated their pain on average over the previous week using an 11-point NRS [25]. Participants were considered to have a previous history of LBP if they answered “yes” to the question: “Have you experienced low back pain in the past”?

#### 2.2.7. Demographics

Age and sex were collected from all participants on the day of baseline testing.

### 2.3. Primary and secondary outcomes recorded at six-month follow-up

The primary outcome was average pain intensity over the past week, assessed using the NRS at six-month follow-up. The secondary outcome was disability assessed using the 24-point Roland Morris Disability Questionnaire (RMDQ) at six-month follow-up [89]. The RMDQ questionnaire detects the level of disability experienced due to LBP. An item receives a score of 1 if it is applicable to the respondent or 0 if it is not, with a total range of 0 (no disability) to 24 (severe disability) [88]. The primary and secondary outcomes were combined to distinguish between those who did and did not report LBP at six months, defined as an NRS score ≥ 2 on average over the previous one-week or ≥ 7 on the RMDQ scale at six-month follow-up [45;108]. This dichotomised outcome measure was used in the secondary analysis, described in detail below.

### 2.4. Sample size

Sample size was calculated *‘a priori’* based on the assumption that at least five variables would show no association with the outcome and be excluded from analysis, and 20% of participants would be lost to follow up. Therefore, 120 participants were required to ensure at least ten subjects per variable within linear regression models (95). Sample size for the logistic regression model was calculated using the rule of thumb that five events per variable are required for adequate statistical power (96). There is substantial variability in the clinical course of acute LBP with estimates for the risk of developing chronic LBP reportedly as high as 56% [97]. Further, recurrence of LBP is common, with 12-month recurrence rates reported in the literature ranging from 24% to 80% [81;99]. This variability suggests a sample size of 120 participants with acute LBP should also be adequate to power the logistic regression analysis (i.e. presence of LBP in 50 or more participants at six months).

### 2.5. Statistical Analyses

Continuous data are presented as mean ± standard deviation (SD) and categorical data presented as percent (%). Statistical significance was accepted at *P* ≤ 0.05. All analyses were conducted with the statistical programming language R, version 4.0.3 (R Development Core Team, Vienna, Austria) [27]. All predictors except sex, previous history of LBP and BDNF genotype were treated as continuous variables. The distribution of the fifteen candidate predictors and their linearity with the outcome variable was inspected using scatter plots. Non-normally distributed variables were log transformed prior to further statistical analyses.

Predictors were assessed for the presence of collinearity as this can produce misleading results and increased standard errors [36]. Spearman’s correlations were calculated between candidate predictors with theoretically plausible associations. Specifically, correlations were calculated between the log transformed N_80_, N_150_ and P_260_ SEP area variables and L3 and L5 map volume.

For the remaining variables, we assumed missing data occurred at random depending on the candidate predictors measured at baseline and the development of future LBP. All missing data were imputed using the multiple imputation by chained equations (MICE) procedure [114]. All available data, including the outcome variables, were used in this procedure [114] to generate thirty imputed data sets. Where data are missing at random (i.e. missing randomly, conditional on covariates), estimates based on multiple imputation are unbiased [58].

Candidate predictors then underwent a variable selection procedure using the least absolute shrinkage and selection operator (lasso) technique [107]. This is a variation from the procedures described in the published study protocol [51]. Penalized regression is the preferred variable selection procedure and decreases the likelihood of overfitting a prediction model [86]. The optimal lasso penalty λ was chosen for each imputed data set using ten-fold cross-validation based on the deviance. To obtain parsimonious models, λ was chosen to be one standard error larger than the optimal λ [106]. This procedure shrinks some coefficients to zero, selecting a subset of predictors in each imputed dataset. The frequency with which each predictor was selected across all datasets was calculated, and a series of candidate models with increasing numbers of included predictors generated. The model displaying the smallest Akaike’s Information Criterion (AIC), across all imputed datasets, was then selected as the final prediction model.

For the primary analysis, outcome variables of pain and disability were treated as continuous data. When an outcome variable is dichotomized, up to one-third of the data is lost [90], causing a loss of statistical power to detect real associations. For a complex and highly individualised construct such as recovery from LBP [48], it is essential to report data as completely as possible. Further, it is of benefit for clinicians and researchers to be aware of the continuous data underpinning commonly reported clinical categories (e.g. chronic LBP). The results of the primary analyses are presented using hierarchical linear regression. Goodness of fit for the linear models was assessed using the R^2^ and adjusted R^2^ value.

Despite the known consequences of forcing continuous data into two groups [1;6;26;49;71], dichotomizing outcome data in clinical research remains widespread [31]. Healthcare providers frequently group information into discrete categories, and for a prognostic model predicting LBP outcome to be useful, comparison with other currently utilised models is required. Therefore, in the secondary analysis the variable selection procedure was repeated using logistic regression to determine predictors of chronic LBP (dichotomized outcome: NRS ≥ 2 or RMDQ ≥ 7).

Predictive discrimination of the model was assessed with the c-statistic (i.e. area under the receiver operating characteristic curve). Model calibration was estimated using Brier Scores [12]. Brier scores range from 0 to 1, with scores closer to 0 (and similar reliability and resolution upon decomposition) representing better calibration. Reliability measures how close the predicted probabilities are to the true probabilities, whereas resolution measures how much the conditional probabilities differ from the prediction average [2]. The presence of miscalibration was further assessed using the Hosmer-Lemeshow test [47]. Despite the known limitations of the Hosmer-Lemeshow test [115], when only internal validation is conducted, overall calibration is guaranteed to be good for logistic regression [102] and calibration-in-the-large becomes irrelevant since the average predicted risks match the event rate [115]. Therefore, it was most appropriate within the current dataset to evaluate calibration numerically using the Brier score and Hosmer-Lemeshow test.

Models predicting dichotomized outcomes perform optimistically within their derived sample [103]. We subjected the prediction model to a ten-fold cross-validation procedure to internally validate our findings [86;103]. This involved splitting the sample ten-fold, with all previously specified imputation and variable selection procedures repeated in 90% of the sample. The predicted probabilities obtained from 90% of the sample are then tested against the outcome in the remaining 10% of the sample. This process is repeated ten times with each subsequent fold, and measures of performance pooled across all imputed, cross-validated datasets with Rubin’s rules.

## 3. RESULTS

### 3.1. Participant characteristics

Of 498 participants who presented with an acute episode of LBP for screening, 120 were included in the study sample (**Figure 2**). Two hundred and seven participants (41.5%) were ineligible because they had chronic LBP, two were ineligible because they had previous spinal surgery and three were ineligible because physical examination by the study investigator suggested a diagnosis of lumbosacral radiculopathy. Of the 286 eligible participants, 94 (32.9%) failed to respond to contact attempts to organise baseline assessment and 72 (25.2%) declined participation after reviewing the study information sheet. Baseline data were obtained on average 2.4 weeks (SD 1.4, range 1 day to 6 weeks) after the onset of an acute LBP episode. Seventy-two participants (60%) were coded GG and 48 (40%) were coded AA/AG for BDNF genotype. According to the Hardy–Weinberg Equilibrium, this observed distribution does not differ significantly from the expected rate (χ^2^ = 1.27, df = 1, *P* = 0.26). Missing data for candidate predictors is presented in **Supplemental File 1**.

**Figure 2.**
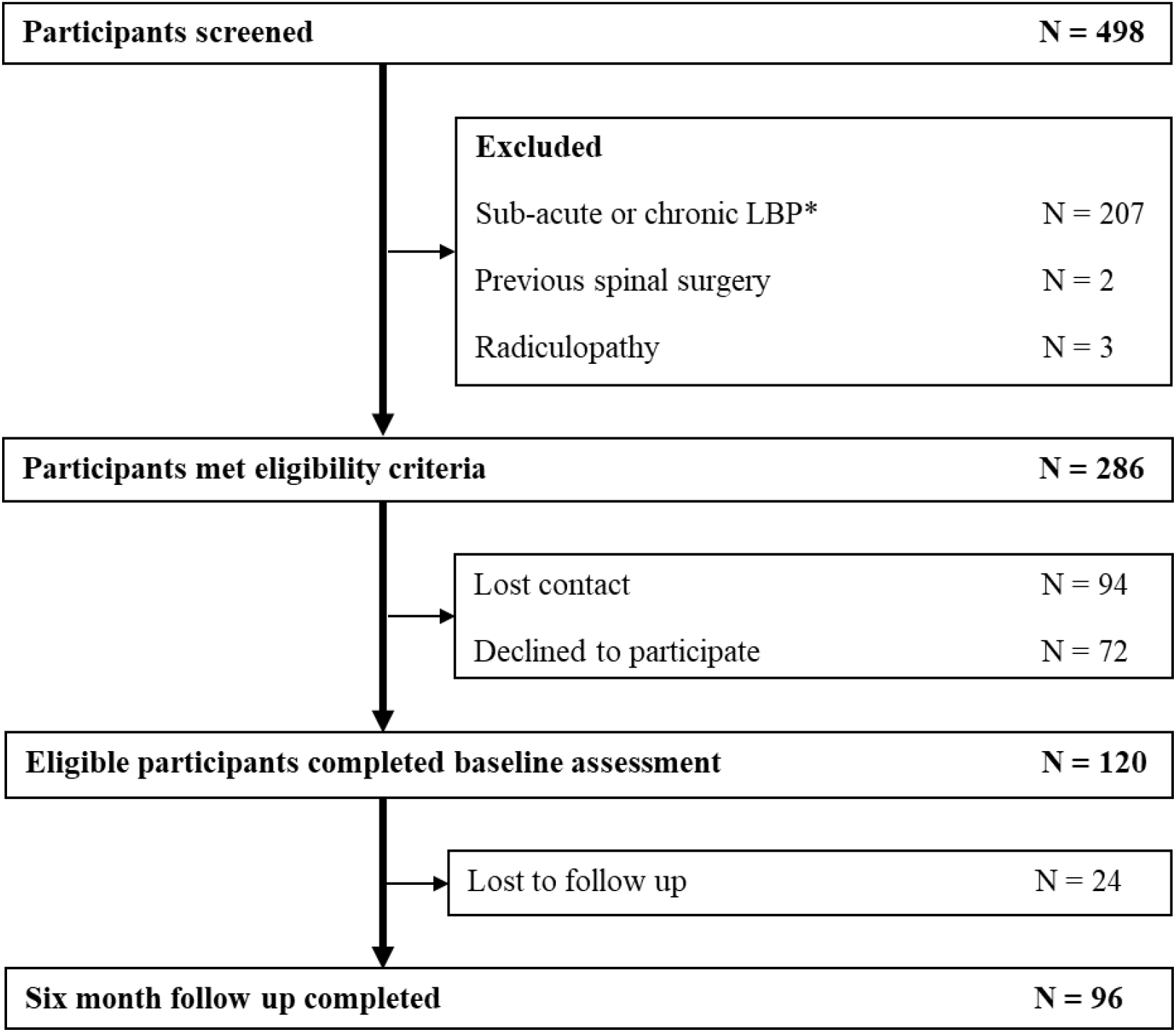
Study flow chart. *defined as LBP lasting for longer than 6 weeks and/or an LBP episode preceded by a period of less than one-month without pain.

Follow-up at six months was completed in 96 participants (80%). Missing follow-up cases were due to the participant failing to respond to multiple contact attempts. The intention was to determine the presence of LBP at six-months (183 days), in practice, follow-up occurred a mean 194 (SD = 20) days after entering the study with an acute episode of LBP. Characteristics of the participants at baseline and univariable associations with chronic LBP are provided in **Table** Individual participant data for normalised motor cortical maps and sensory evoked potentials are displayed in **Figure 3 and 4**.

**Figure 3.**
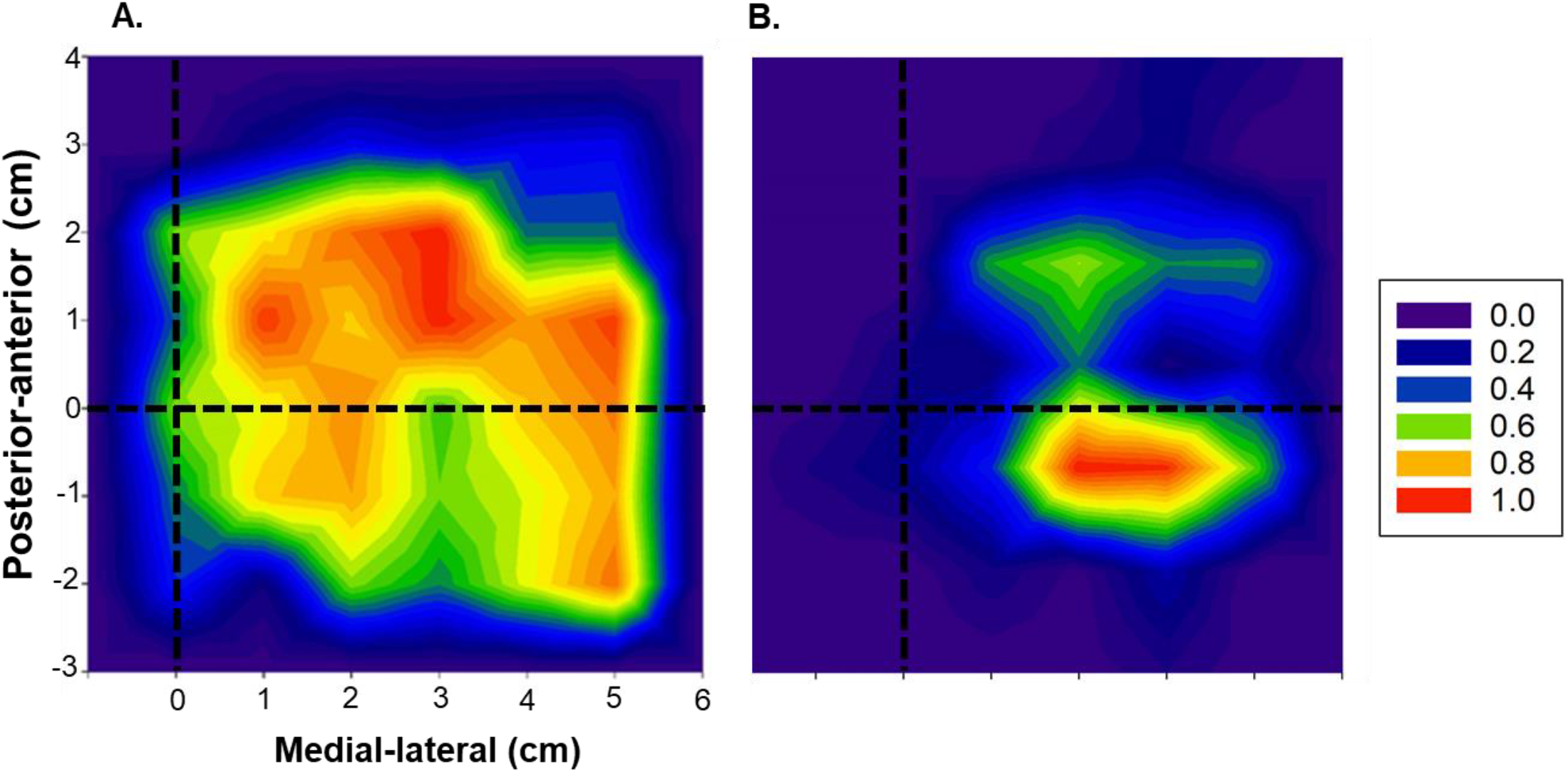
Motor cortical maps for two representative participants at the L3 recording site normalized to peak motor evoked potential (MEP) amplitude. **Figure 3A** displays large volume corticomotor excitability during acute low back pain (LBP) in a participant who was recovered at 6-months. **Figure 3B** displays small volume corticomotor excitability during acute LBP in another participant who experienced chronic/recurrent LBP at 6-months. The dashed lines indicate the location of the vertex (coordinate 0,0). The coloured scale represents the proportion of the maximum MEP amplitude. Warmer colours represent higher excitability.

**Figure 4.**
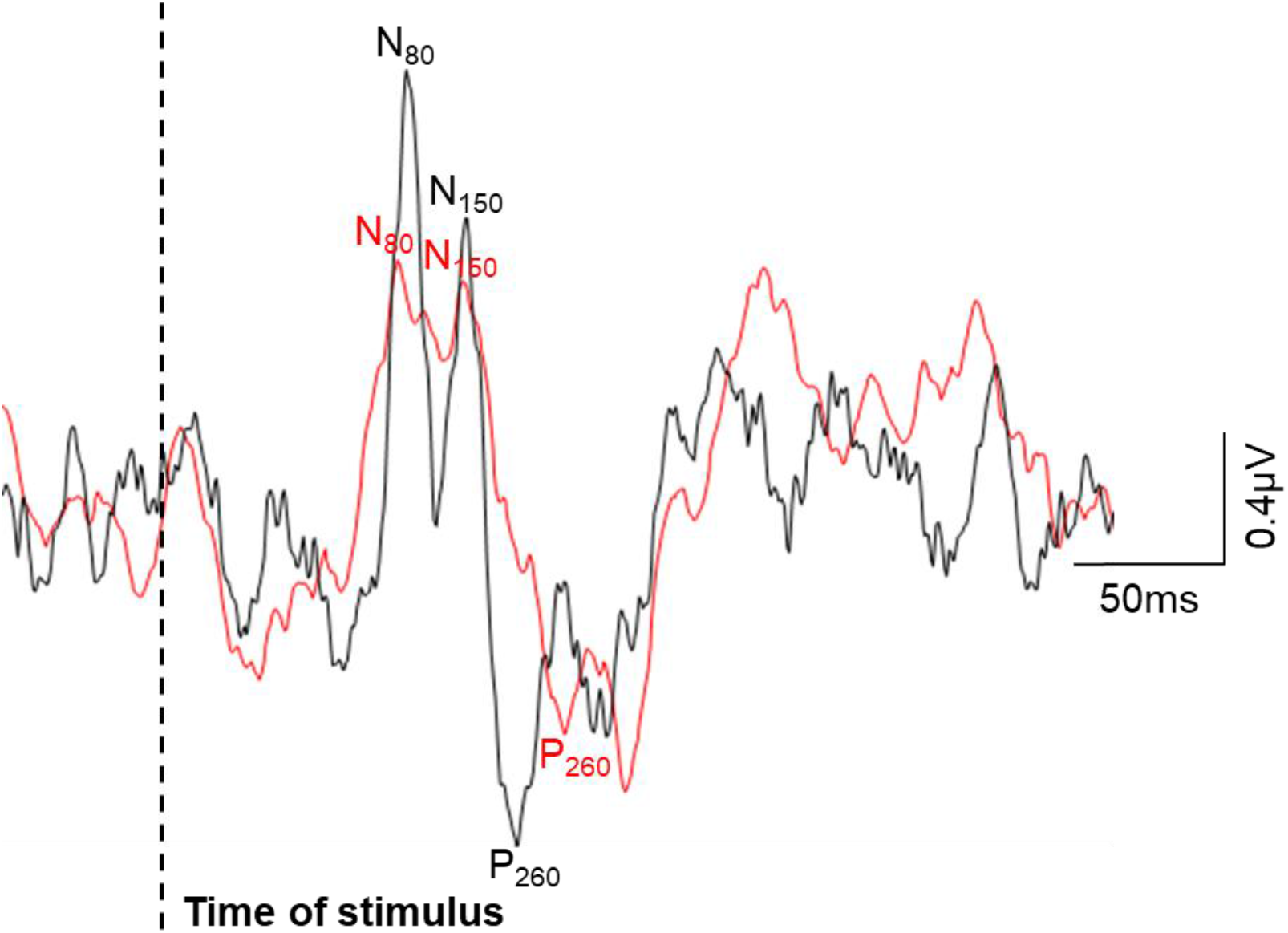
Sensory evoked potentials (SEP) recorded in response to stimuli to the paraspinal muscles at baseline (average of approx. 500 traces) for two representative participants. The black trace displays high SEP excitability in the acute stage of low back pain (LBP) in a participant who was recovered at 6-months. The red trace displays low sensory evoked potential excitability in the acute stage of LBP in a participant who reported LBP at 6-months.

### 3.2. Incidence of chronic LBP

Analysis of complete cases revealed that 54% of participants reported LBP at six-month follow-up (average pain intensity 3.9 [SD = 1.7], average RMDQ score 4.8 [SD = 5.4]) and could be considered to have persistent or recurring LBP. This is comparable to the incidence of chronicity in previous Australian estimates [42]. Of the 52 participants deemed to have LBP, only 12 participants (23%) had an RMDQ score ≥ 7. The remaining 46% of participants were classified as recovered (average pain intensity 0.3 [SD = 0.5], average RMDQ score 0.9 [SD = 1.6]).

### 3.3. Predictor selection before modelling

All variables were normally distributed except the N_80_, N_150_ and P_260_ SEP area and these variables were log-transformed. Baseline characteristics were then compared between participants who did (N=96), and did not (N=24), complete follow up at six months (**Table 2**). Apart from N_150_ and P_260_ SEP area measures, no statistically significant differences at baseline were identified between participants who did, or did not, complete follow-up, however, the N_80_ SEP area did demonstrate a strong tendency (*P* = 0.06). Next, spearman’s correlation coefficients were calculated between all measures of cortical excitability (**Table 3**). No strong correlation was identified between SEP and TMS measures. The N_80_ and N_150_ (r_s_ = 0.84, P = < 0.001), and N_80_ and P_260_ (r_s_ = 0.85, P = <0.001) SEP area values were strongly correlated. As the N_150_ and P_260_ SEP area measurements may have impacted the missing at random assumption and demonstrated collinearity with the N_80_ SEP area they were excluded from further analyses. The remaining thirteen candidate predictors were subjected to the λ-1se variable selection procedure.

**Table 1.**
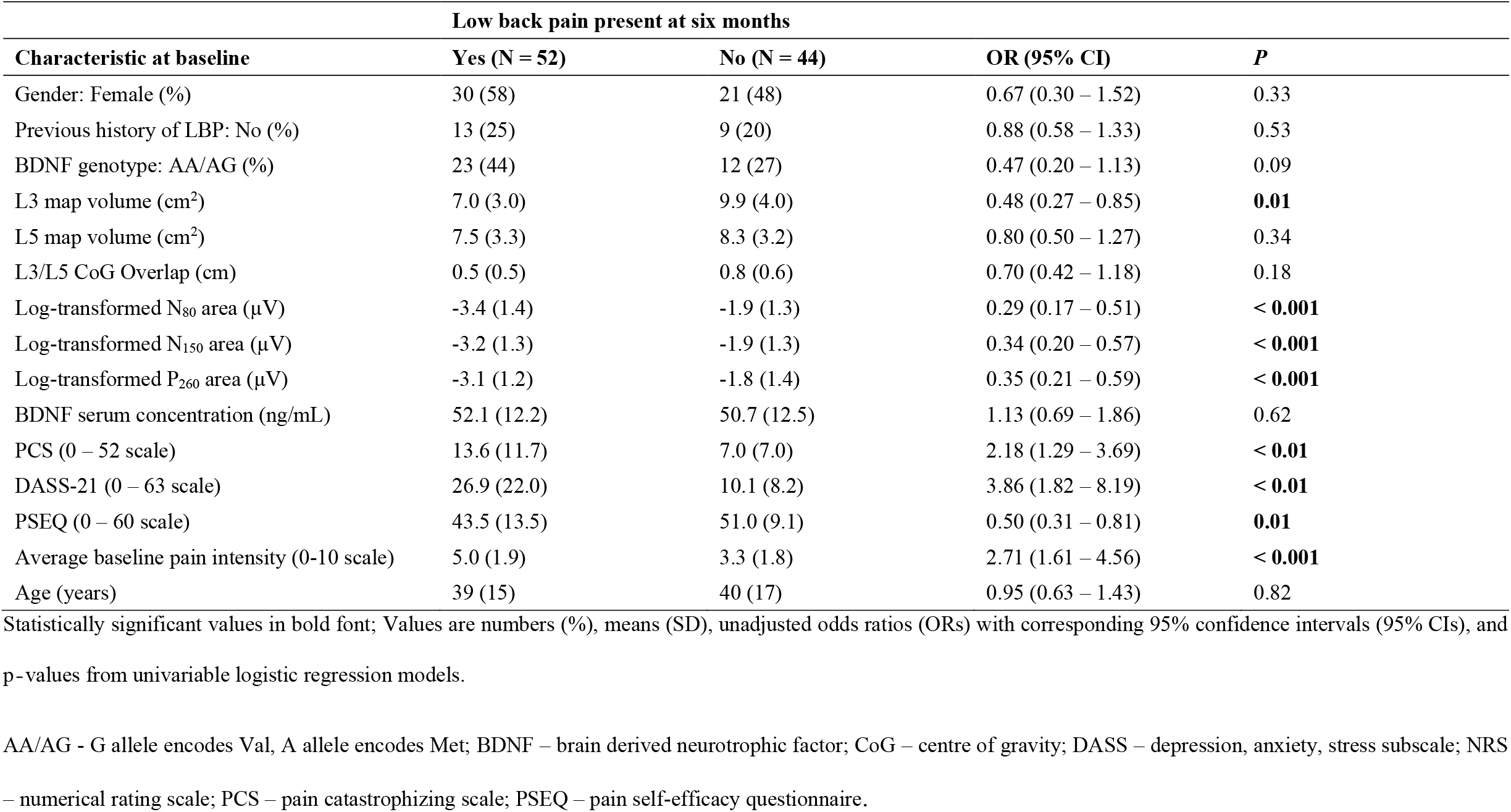
Baseline characteristics of participants who completed six-month follow-up, with (N = 52) or without (N = 44) low back pain (LBP) at six months.

| Characteristic at baseline | Low back pain present at six months |  | OR (95% CI) | P |
| --- | --- | --- | --- | --- |
|  | Yes (N = 52) | No (N = 44) |  |  |
| Gender: Female (%) | 30 (58) | 21 (48) | 0.67 (0.30 – 1.52) | 0.33 |
| Previous history of LBP: No (%) | 13 (25) | 9 (20) | 0.88 (0.58 – 1.33) | 0.53 |
| BDNF genotype: AA/AG (%) | 23 (44) | 12 (27) | 0.47 (0.20 – 1.13) | 0.09 |
| L3 map volume (cm<sup>2</sup>) | 7.0 (3.0) | 9.9 (4.0) | 0.48 (0.27 – 0.85) | **0.01** |
| L5 map volume (cm<sup>2</sup>) | 7.5 (3.3) | 8.3 (3.2) | 0.80 (0.50 – 1.27) | 0.34 |
| L3/L5 CoG Overlap (cm) | 0.5 (0.5) | 0.8 (0.6) | 0.70 (0.42 – 1.18) | 0.18 |
| Log-transformed N<sub>80</sub> area (μV) | -3.4 (1.4) | -1.9 (1.3) | 0.29 (0.17 – 0.51) | **< 0.001** |
| Log-transformed N<sub>150</sub> area (μV) | -3.2 (1.3) | -1.9 (1.3) | 0.34 (0.20 – 0.57) | **< 0.001** |
| Log-transformed P<sub>260</sub> area (μV) | -3.1 (1.2) | -1.8 (1.4) | 0.35 (0.21 – 0.59) | **< 0.001** |
| BDNF serum concentration (ng/mL) | 52.1 (12.2) | 50.7 (12.5) | 1.13 (0.69 – 1.86) | 0.62 |
| PCS (0 – 52 scale) | 13.6 (11.7) | 7.0 (7.0) | 2.18 (1.29 – 3.69) | **< 0.01** |
| DASS-21 (0 – 63 scale) | 26.9 (22.0) | 10.1 (8.2) | 3.86 (1.82 – 8.19) | **< 0.01** |
| PSEQ (0 – 60 scale) | 43.5 (13.5) | 51.0 (9.1) | 0.50 (0.31 – 0.81) | **0.01** |
| Average baseline pain intensity (0-10 scale) | 5.0 (1.9) | 3.3 (1.8) | 2.71 (1.61 – 4.56) | **< 0.001** |
| Age (years) | 39 (15) | 40 (17) | 0.95 (0.63 – 1.43) | 0.82 |

Statistically significant values in bold font; Values are numbers (%), means (SD), unadjusted odds ratios (ORs) with corresponding 95% confidence intervals (95% CIs), and p-values from univariable logistic regression models.
AA/AG - G allele encodes Val, A allele encodes Met; BDNF – brain derived neurotrophic factor; CoG – centre of gravity; DASS – depression, anxiety, stress subscale; NRS – numerical rating scale; PCS – pain catastrophizing scale; PSEQ – pain self-efficacy questionnaire.

**Table 2.** Comparison of baseline characteristics between participants who did, and did not, complete six-month follow-up.

| Characteristic | Completed follow-up (N = 96) | Did not complete follow-up (N = 24) | P-value |
| --- | --- | --- | --- |
| Gender: Female (%) | 53.1 | 33.3 | 0.08 |
| Previous history of LBP: No (%) | 22.9 | 12.5 | 0.32 |
| BDNF genotype: AA/AG (%) | 36.5 | 54.2 | 0.11 |
| L3 map volume (cm<sup>2</sup>) | 8.4 (4.0) | 10.0 (5.5) | 0.13 |
| L5 map volume (cm<sup>2</sup>) | 7.8 (3.5) | 8.5 (4.2) | 0.43 |
| L3/L5 CoG Overlap (cm) | 0.7 (0.6) | 0.5 (0.4) | 0.21 |
| Log-transformed N<sub>80</sub> area (μV) | -1.2 (0.7) | -1.5 (0.6) | 0.06 |
| Log-transformed N<sub>150</sub> area (μV) | -1.1 (0.6) | -1.4 (0.6) | **0.04** |
| Log-transformed P<sub>260</sub> area (μV) | -1.1 (0.6) | -1.4 (0.6) | **0.03** |
| BDNF serum concentration (ng/mL) | 52.2 (13.7) | 50.9 (13.7) | 0.71 |
| PCS (0 – 52 scale) | 10.6 (10.3) | 12.6 (8.9) | 0.35 |
| DASS-21 (0 – 63 scale) | 19.1 (19.3) | 27.2 (24.7) | 0.17 |
| PSEQ (0 – 60 scale) | 47.1 (12.1) | 42.5 (11.5) | 0.10 |
| Average baseline pain intensity (0-10 scale) | 4.2 (2.0) | 4.6 (1.3) | 0.37 |
| Age (years) | 40 (16) | 40 (12) | 0.91 |

Statistically significant values in bold font
AA/AG - G allele encodes Val, A allele encodes Met; BDNF – brain derived neurotrophic factor; CoG – centre of gravity; DASS – depression, anxiety, stress subscale; NRS – numerical rating scale; LBP – low back pain; PCS – pain catastrophizing scale; PSEQ – pain self-efficacy questionnaire.

**Table 3.**
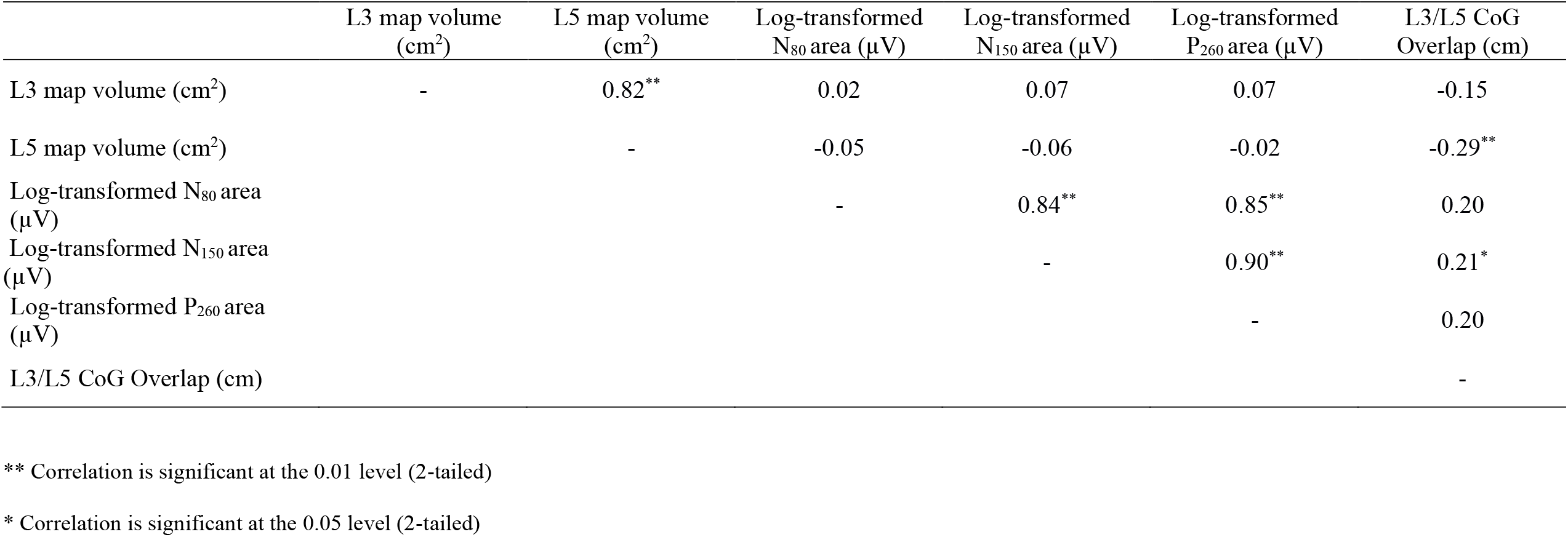
Spearman’s correlation coefficients between measures of cortical excitability during acute low back pain.

### 3.4. Predictors of pain intensity at six months

Higher baseline pain intensity (*P* = <0.001), higher depression and anxiety (DASS-21: *P* = 0.31), higher pain catastrophizing (PCS: P = 0.39), lower N_80_ SEP area (*P* = 0.01) and lower L3 map volume (*P* = < 0.01) predicted worse pain over the past week (continuous outcome) at six months. The results of hierarchical regression modelling are presented in **Table 4**. Baseline pain intensity explained 22% of the variance in pain intensity at six-months. Addition of the DASS-21 and PCS instruments explained a further 10% of the variance (F_[2,88]_ = 4.99, *P* = 0.01) and the addition of neurobiological predictors (N_80_ SEP area and L3 map volume) explained a further 15% of the variance in six-month pain intensity (F_[2,76]_ = 8.61, *P* = < 0.001). In combination, these five variables explained 47% (R^2^ = 0.47 95% CI = 0.31 – 0.60) of the total variance in six-month pain intensity.

**Table 4.** Hierarchical linear regression model predicting six-month pain intensity.

| Variables entered |  | R<sup>2</sup> | Adjusted R<sup>2</sup> | Significance of<br>Adjusted R<sup>2</sup> change | B (95% CI)<br>(final model) | Significance of B |
| --- | --- | --- | --- | --- | --- | --- |
| Step 1 | Average baseline pain intensity (0-10 scale) | 0.22 | 0.22 |  | 0.33 (0.17 to 0.48) | < **0.001** |
| Step 2 | DASS-21 (0 – 63 scale) |  |  |  | 0.14 (-0.13 to 0.41) | 0.31 |
|  | PCS (0 – 52 scale) | 0.32 | 0.30 | **0.01** | 0.10 (-0.13 to 0.35) | 0.39 |
| Step 3 | L3 map volume (cm<sup>2</sup>) |  |  |  | -0.29 (-0.48 to -0.10) | < **0.01** |
|  | Log-transformed N<sub>80</sub> area (μV) | 0.47 | 0.44 | < **0.01** | -0.25 (-0.42 to -0.07) | **0.01** |

Statistically significant *P*-values in bold font
Hierarchical linear regression was performed with six-month pain intensity as the dependent variable. Predictors identified from the λ-1se variable selection procedure were entered into the model. Step 1 included average baseline pain intensity, step 2 included psychological risk factors and step 3 included measures of sensorimotor cortical excitability. The significance of the adjusted R<sup>2</sup> change was calculated using a one-way ANOVA.
B – unstandardized beta coefficient; CI – confidence interval; DASS-21 – 21 item depression, anxiety, stress subscale; PCS – pain catastrophizing scale

### 3.5. Predictors of disability at six months

Higher pain catastrophizing (PCS: B = 0.47, 95% CI = 0.30 to 0.64, *P* = < 0.001) and older age (B = 0.24, 95% CI = 0.08 to 0.42, *P* = < 0.01) predicted greater levels of disability over the past week (continuous outcome) at six months. These two variables explained 30% of the variance in disability outcome at six months (R^2^ = 0.30, 95% CI = 0.14 to 0.46).

### 3.6. Predictors of presence or not of LBP at six months

When the dichotomous variable of LBP at six-months was designated on the basis of pain or disability above a threshold value (NRS ≥ 2 or RMDQ ≥ 7), six predictors remained in the multivariable logistic regression model following λ-1se variable selection and ten-fold cross-validation **(Table 5)**. Lower primary sensory cortex excitability (N_80_ SEP area: P = < 0.01), lower corticomotor excitability (L3 map volume: P = 0.07), BDNF genotype MET allele carriers (P = 0.22), higher depression and anxiety (DASS-21 score: P = 0.01), no prior history of LBP (P = 0.15) and higher baseline pain intensity (P = 0.01) predicted the presence of LBP at six-months.

**Table 5.** Multivariate logistic prediction model of risk of low back pain at six months.

| Predictor | B | Odds Ratio (95% CI) | Significance of B |
| --- | --- | --- | --- |
| Log-transformed N<sub>80</sub> area (µV) | -0.79 | 0.45 (0.27 to 0.75) | < **0.01** |
| L3 map volume (cm<sup>2</sup>) | -0.21 | 0.81 (0.65 to 1.01) | 0.07 |
| BDNF genotype (0 = AA/AG, 1 = GG) | -0.87 | 0.41 (0.10 to 1.69) | 0.22 |
| DASS-21 (0 – 63 scale) | 0.07 | 1.07 (1.02 to 1.23) | **0.01** |
| Average baseline pain intensity (0-10 scale) | 0.54 | 1.71 (1.17 to 2.50) | **0.01** |
| Previous history of LBP (0 = no, 1 = yes) | -1.29 | 0.27 (0.05 – 1.61) | 0.15 |

Statistically significant values in bold font
AA/AG - G allele encodes Val, A allele encodes Met; B – unstandardized beta coefficient; BDNF – brain derived neurotrophic factor; CI – confidence interval; DASS-21 – 21 item depression, anxiety, stress subscale; GG – G allele encodes Val; LBP – low back pain.

The logistic prediction model demonstrated outstanding discrimination. The c-statistic was 0.91 (95% CI 0.84 to 0.95) indicating that for 91% of randomly chosen pairs of study participants, in which one reported LBP at six months and the other did not, the participant with the higher predicted risk was the one who reported LBP. The Brier Score was 0.12 (SD = 0.03) and decomposition of the Brier Score revealed reliability of 0.03 (SD = 0.02) and resolution of 0.15 (SD = 0.02). The Hosmer-Lemeshow test was not statistically significant (*P* = 0.35), thus there was no evidence of model miscalibration.

A simplified model considering only the three statistically significant predictors of LBP at six-months (i.e. N_80_ SEP area, DASS-21, baseline pain intensity) also demonstrated excellent predictive performance with a c-statistic of 0.86 (95% CI 0.79 to 0.92) and acceptable calibration (Brier Score = 0.15 [SD = 0.02], reliability = 0.01 [SD = 0.01], resolution = 0.10 [SD = 0.02], Hosmer-Lemeshow test [P = 0.18]).

### 3.7. Sensitivity analysis

A sensitivity analysis was conducted to explore the effect of imputing missing six-month outcome data on study results. Participants who did not return for follow-up at six months were removed from the dataset and all modelling steps repeated. Predictors retained in each model following λ-1se variable selection were identical for all outcomes. The multivariable linear model predicting six-month pain intensity had a R^2^ value of 0.45 (95% CI = 0.29 – 0.60) and the linear model predicting six-month disability score had a R^2^ value of 0.32 (95% CI = 0.17 – 0.47). Predictive performance of the six variable logistic prediction model was also comparable, demonstrating a c-statistic of 0.88 (95% CI = 0.81 to 0.95). The Hosmer-Lemeshow test was not statistically significant (*P* = 0.59) and the Brier Score was 0.13 [SD=0.02]) suggesting comparable calibration.

## 4. DISCUSSION

This prospective longitudinal cohort study is the first to integrate biological measures of sensorimotor cortical function and neuroplasticity, psychological, symptom-related, and demographic factors in a prognostic model of the long-term outcome after an acute episode of LBP. A novel finding was the identification of lower primary sensory cortex excitability (N_80_ SEP area) and lower corticomotor excitability (L3 map volume) in the acute stage of LBP as risk factors for higher pain intensity reported at six months, and in conjunction with the BDNF MET allele, as risk factors for future LBP. When these variables were combined with psychological factors (higher emotional distress) and symptom-related factors (no prior LBP history, higher baseline pain intensity), they explained a similar percentage of the variance in six month pain intensity (47%) as earlier models that integrate psychological and symptom-related factors (46%) [57], but uniquely, the prognostic model accurately predicted the presence of LBP at six months in 91% of individuals. This predictive accuracy is higher than that of similar prognostic models and may assist in the early identification of individuals at risk of persisting or recurring LBP.

The primary analysis aimed to identify the strongest predictors of pain intensity and disability at six months as continuous outcomes. These analyses revealed largely discrete risk factors for the outcomes of pain (strongest risk factors of lower sensory cortex excitability [N_80_ SEP area], lower corticomotor excitability [L3 map volume], higher baseline pain intensity with a lesser contribution from higher emotional distress and higher pain catastrophising) and disability (older age, higher pain catastrophizing), with no biological risk factors identified for disability. Patient-reported outcomes of pain (e.g. NRS) and disability (e.g. RMDQ) are known to be weakly correlated with each other when LBP is chronic [62] and previous research has shown that disability is more closely aligned with psychological risk factors than is pain intensity. For example, in a large cohort study of people with chronic LBP, psychological questionnaires thought to assess ‘pain-related distress’ explained 51% of the variance in disability, but only 35% of the variance in pain intensity [15]. The absence of a relationship between low sensorimotor cortex excitability and disability suggests other factors predict this outcome.

The secondary analysis aimed to develop a prognostic model to identify those at risk of future LBP. Several previous studies have attempted to develop prognostic models in LBP [40]. Those studies have generally focussed on self-report psychosocial and symptom-related factors with low predictive accuracy [33;70;85]. In addition, few have undergone internal and external validation [54]. These limitations hamper the ability of currently available prognostic models to identify those at risk of future LBP.

Our findings on the psychological and symptom-related risk factors for chronic LBP are consistent with prior work [40;68;85]. We confirm previous findings that show that higher emotional distress and higher pain in the acute stage of LBP are risk factors for the development of persistent or recurring LBP. The discovery that lower sensory cortex excitability and lower corticomotor excitability are risk factors for future LBP is novel. When N_80_ SEP area, L3 map volume and BDNF genotype were combined with psychological (higher emotional distress) and symptom-related (no prior LBP history, higher baseline pain intensity) factors, the prognostic model had very high discriminatory performance (c-statistic 0.91 [0.84 to 0.95]). Even when a simplified model inclusive only of N_80_ SEP area, emotional distress and baseline pain intensity was evaluated, discrimination remained high (c-statistic 0.86 [0.79 to 0.92]. These findings suggest that a small N_80_ SEP area, thought to reflect low primary somatosensory cortex excitability, may be an important risk factor for the development of future pain. Further, these findings highlight the importance of assessing diverse phenotypic traits, across a range of neurobiological, psychological, symptom-related and demographic domains, when attempting to predict LBP outcome. Previous studies utilising cluster analyses have shown stronger associations between LBP outcome and psychological factors when relevant biological factors are considered in tandem [60;61]. For example, in a longitudinal study using cluster analyses, individuals with the worst recovery from acute LBP at six-month follow-up displayed higher depression-like symptoms in conjunction with higher serum concentrations of tumor necrosis factor [60].

The discovery that low primary sensory cortex excitability and low corticomotor excitability in the acute stage of LBP are risk factors for chronic LBP builds on a growing body of evidence that suggests measures of brain structure and function can predict future LBP. For example, in sub-acute LBP, greater functional connectivity of corticostriatal circuitry predicts chronic LBP at 1-year [8;72]. Further, using causal inference analyses, we have shown that low sensory cortex excitability (N_80_ SEP area) in acute LBP is a cause, and not an epiphenomenon, of chronic pain [50]. Our findings also demonstrate that BDNF genotype MET allele carriers are more susceptible to future LBP. Together these findings support current theories suggesting chronic pain is a consequence of neuroplastic changes occurring across a distributed network of brain regions that has both heritable and environmental contributions. Although the precise relationship between these predictors and the development of chronic LBP remains to be elucidated, it is plausible that sensorimotor cortex excitability may represent a modifiable risk factor that, if increased in the acute stage of LBP (e.g. using non-invasive brain stimulation, peripheral electrical stimulation or other behavioural interventions), might interfere with the transition to chronic LBP. Further research is needed to explore this possibility.

One argument against the inclusion of neurobiological variables in prognostic models and subsequently, clinical screening tools for chronic LBP, is the feasibility (e.g. expertise and equipment requirements) of implementing these measures in clinical practice. Although expertise and specialised equipment are required, sensory evoked potentials are already used as a diagnostic and prognostic tool in clinical settings for the evaluation of neurological disorders. Electroencephalography recordings can be made in minutes using single scalp electrodes or ‘wash and wear’ caps and the electrical stimulation devices needed to deliver non-noxious stimulation are inexpensive and available in many healthcare settings. Should external validation confirm the importance of lower N_80_ SEP area as a key risk factor for future LBP, detection methods and devices could be rapidly optimised, and key performance characteristics determined, for implementation in clinical practice.

Despite a robust and rigorous approach to data collection and analyses, this study has some limitations. First, multivariable logistic prediction models perform optimistically in the sample in which they are developed. We calibrated our model appropriately and minimized optimism using penalized regression and cross-validation techniques [38;86;103]. However, all prediction models developed in this study require external validation [104]. Second, missing data were present within some candidate predictors and no outcome data were available for 24 participants. To limit the impact of missing data on our findings, missing values were imputed using the MICE procedure [101] and a sensitivity analysis identified no differences between predictions developed with or without imputed outcome data. Third, our outcome measures for pain and disability asked participants to report these features over the past week, and we cannot determine whether LBP had persisted since the acute episode (i.e., defined as chronic) or whether their LBP had reoccurred as a new discrete episode (i.e., defined as recurrent). For this reason, we refer to our results as predictors of future LBP and cannot determine whether risk factors differ between chronic or recurring LBP. Finally, presence of LBP at six months was designated based on both pain and disability above an *a priori* reported threshold at follow-up. The threshold value of ≥ 2 on the NRS or ≥ 7 on the RMDQ scale was chosen as it reflects cut-offs used in previous LBP prognostic research allowing direct comparison of predictions [45;48;108].

### 4.1. Conclusion

This study identified novel risk factors relating to cortical function and neuroplasticity for the development of future LBP that could predict an individual’s pain intensity and level of disability at six-month follow-up, and accurately discriminate between those who did, and did not, have LBP at this time-point. Future research that externally validates these findings may lead to the development of a prognostic model with clinical applicability to identify individuals at high risk of developing chronic LBP.

## Data Availability

The data that support the findings of this study are available from the corresponding author, [SMS], upon reasonable request.

## Acknowledgements

We thank the patients who participated in the study. We thank Dr Valerie Wasinger, Biosciences Precinct Laboratory, Level 2, Biosciences South (E26), UNSW Sydney, NSW 2052 for her expertise and assistance with serum analyses.

