## Supplemental File 1 for "Cortical function and sensorimotor plasticity predict future low back pain after an acute episode: the UPWaRD prospective cohort study"

**Supplemental File 1.** Missing data within candidate predictor variables.

| Candidate Predictor | Number of missing data (%) | Reason |
|---|---|---|
| Somatosensory and anterior cingulate cortex excitability | | |
| SEP $N_{80}$ component area | 2 (2%) | Equipment failure |
| SEP $N_{150}$ component area | 2 (2%) | Equipment failure |
| SEP $P_{260}$ component area | 2 (2%) | Equipment failure |
| Corticomotor excitability | | |
| L3 map volume | 31 (25.8%) | Unresolvable noise to signal ratio (N = 5) |
| L5 map volume | 31 (25.8%) | Consent not obtained (N = 13) |
| L3/L5 centre of gravity overlap | 31 (25.8%) | Participant unable to tolerate (N = 7) |
| | | Equipment failure (N = 6) |
| Markers of neuroplastic potential | | |
| BDNF genotype | 0 (0%) | N/A |
| BDNF serum concentration | 30 (25%) | Researcher error during phlebotomy (N = 10) |
| | | Consent not obtained (N = 11) |
| | | Simple plex Ella™ machine error (N = 9) |
| Psychological status | | |
| PCS | 2 (2%) | Incorrect completion of questionnaire |
| DASS-21 | 7 (5.8%) | Incorrect completion of questionnaire |
| PSEQ | 3 (2.5%) | Incorrect completion of questionnaire |
| Symptom-related factors | | |
| NRS score at T1 | 2 (2%) | Incorrect completion of questionnaire |
| Previous history of low back pain | 4 (3%) | Incorrect completion of questionnaire |
| Demographics | | |
| Age | 0 (0%) | N/A |
| Sex | 0 (0%) | N/A |

BDNF, brain-derived neurotrophic factor; DASS-21, Depression, Anxiety and Stress Scale; L3, electrode recording site 3cm lateral to the L3 spinous process; L5, electrode recording site 1cm lateral to the L5 spinous process; PCS, Pain Catastrophising Scale; PSEQ, Pain Self-Efficacy Questionnaire; SEP, sensory evoked potential; T1, within 6 weeks of acute low back pain onset; NRS, 11-point numerical rating scale.
